# Standardization of enzyme-linked immunosorbent assays for serosurveys of the SARS-CoV-2 pandemic using clinical and at-home blood sampling

**DOI:** 10.1101/2020.05.21.20109280

**Authors:** Carleen Klumpp-Thomas, Heather Kalish, Matthew Drew, Sally Hunsberger, Kelly Snead, Michael P Fay, Jennifer Mehalko, Anandakumar Shunmugavel, Vanessa Wall, Peter Frank, John-Paul Denson, Min Hong, Gulcin Gulten, Simon Messing, Jennifer Hicks, Sam Michael, William Gillette, Matthew D Hall, Matthew Memoli, Dominic Esposito, Kaitlyn Sadtler

## Abstract

The extent of SARS-CoV-2 infection throughout the United States population is currently unknown. High quality serology is a key tool to understanding the spread of infection, immunity against the virus, and correlates of protection. Limited validation and testing of serology assays used for serosurveys can lead to unreliable or misleading data, and clinical testing using such unvalidated assays can lead to medically costly diagnostic errors and improperly informed public health decisions. Estimating prevalence and clinical decision making is highly dependent on specificity. Here, we present an optimized ELISA-based serology protocol from antigen production to data analysis. This protocol defines thresholds for IgG and IgM for determination of seropositivity with estimated specificity well above 99%. Validation was performed using both traditionally collected serum and dried blood on mail-in blood sampling kits, using archival (pre-2019) negative controls and known PCR-diagnosed positive patient controls. Minimal cross-reactivity was observed for the spike proteins of MERS, SARS1, OC43 and HKU1 viruses and no cross reactivity was observed with anti-influenza A H1N1 HAI titer during validation. This strategy is highly specific and is designed to provide good estimates of seroprevalence of SARS-CoV-2 seropositivity in a population, providing specific and reliable data from serosurveys and clinical testing which can be used to better evaluate and understand SARS-CoV-2 immunity and correlates of protection.

## INTRODUCTION

SARS-CoV-2 has spread across the globe rapidly causing a worldwide pandemic^1^. Infection with this highly contagious respiratory virus can be asymptomatic or present as COVID19, a disease with varying levels of severity that includes a broad range of not fully understood symptoms that may include fever, cough, anosmia, gastrointestinal symptoms, hypercoagulability, inflammatory complications, acute respiratory distress syndrome (ARDS), as well as death^2,3^. Due to the rapidly evolving nature of pandemics, the true extent of spread of SARS-CoV-2 will likely not fully be realized until late or after the pandemic. Moreover, as observed in all respiratory viral pandemics since 1918, the true number of infections always exceeds the detected cases^4,5^. In order to determine a better estimate of the prevalence of SARS-CoV-2 infection, high quality serology assays must be developed. These assays measure the presence of antibodies against specific proteins of this novel coronavirus to determine whether an individual has been infected with SARS-CoV-2, and aim for high sensitivity and specificity^6,7^. Both are important factors to clinically diagnose prior infection; however, if a tradeoff between sensitivity and specificity is needed, high specificity should be emphasized when determining the extent of exposure across a population or for clinically diagnosing previous infections. If such a highly specific, high quality assay is available then data can be generated from serosurveys and clinical testing that can be used to better understand spread of infection, immunity, and correlates of protection.

In order to properly prepare to generate such useful data from an ongoing National Institutes of Health (NIH) sponsored national serosurvey in the United States (NCT04334954) we have developed a serology protocol that emphasizes specificity while maintaining a simple approach that can be repeated at relatively low cost in labs without specialized equipment. The NIH serosurvey study allows mail-in home sampling using dried blood on a microsampler or collection of blood on-site. Therefore we developed, implemented, and evaluated a serology testing protocol using enzyme-linked immunosorbent assays (ELISA) that could successfully be used with multiple sample types while emphasizing the necessary specificity required to conduct high quality convalescent testing and serosurveys (**Figure 1**). Here we present an optimized ELISA-based serology assay protocol -- from protein production to data analysis -- that analyzes the presence of IgG, IgM, and IgA antibodies against spike and RBD antigens of the SARS-CoV-2 coronavirus. This protocol includes: steps for determining the possibility of cross reactivity against a panel of spike antigens from other beta-coronaviruses, control samples, and criteria for setting threshold cut points. A semi-automated protocol is also described that significantly increases throughput capacity.

**Figure 1:**
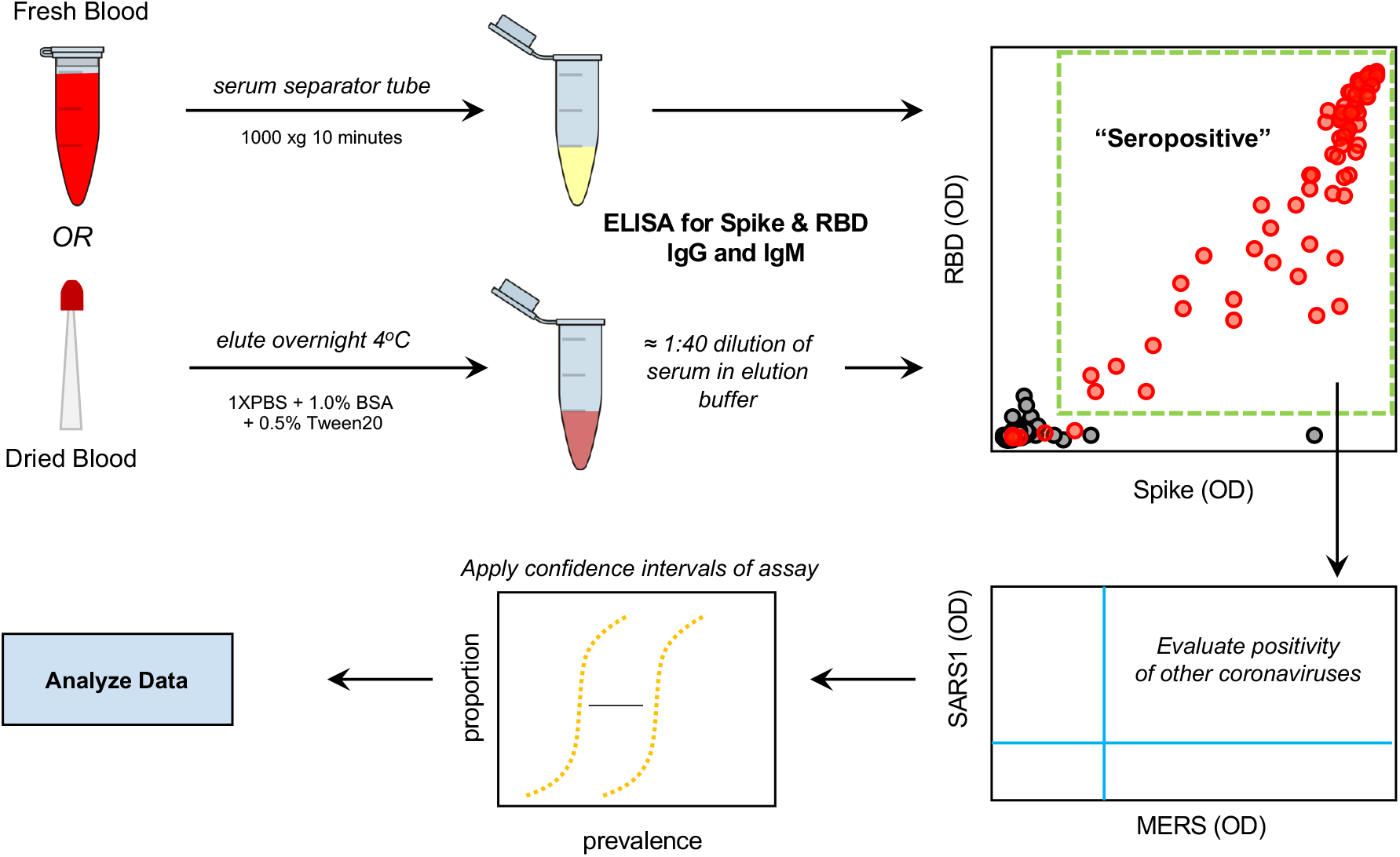
Serology testing protocol for evaluation of SARS-CoV-2 seropositivity in a large-scale population. Utilizing both venipuncture-derived fresh blood and dried blood spots, we have standardized a dual-antigen ELISA platform for highly specific (IgG = 100%, 95% confidence interval = 98.5 – 100) detection of SARS-CoV-2 antibodies for application in precise, large-scale serosurvey efforts.

## MATERIALS AND METHODS

### Cloning and DNA preparation

DNA for the expression of VRC spike (VRC-SARS-CoV-2 S-2P-3C-His8-Strep2x2) and spike proteins for SARS-CoV, MERS-CoV, OC43-CoV, and HKU1-CoV were generously provided by Dr. Kizzmekia Corbett (VRC, NIAID). DNA for the expression of Mt Sinai spike (Kram-SARS-CoV-2 S-2P-His6) and Mt. Sinai RBD (Kram-SARS-CoV-2 S-RBD(319-541)-His6) were generously provided by Dr. Florian Krammer (Mt. Sinai School of Medicine) through BEI Resources. DNA for the expression of Ragon RBD (Ragon-SARS-CoV-2 S-RBD(319-529)-3C-His8-SBP) was generously provided by Dr. Aaron Schmidt (Ragon Institute of MGH, MIT, and Harvard, Norman et al *medRxiv* 2020). Schematics of the similarities and differences in these constructs are shown in **Figure 2a**. Transfection-quality DNA was produced inhouse using the Qiagen Plasmid Plus Maxi Kit per the manufacturer’s protocols or was generated at large-scale by Aldevron (Fargo, ND).

**Figure 2:**
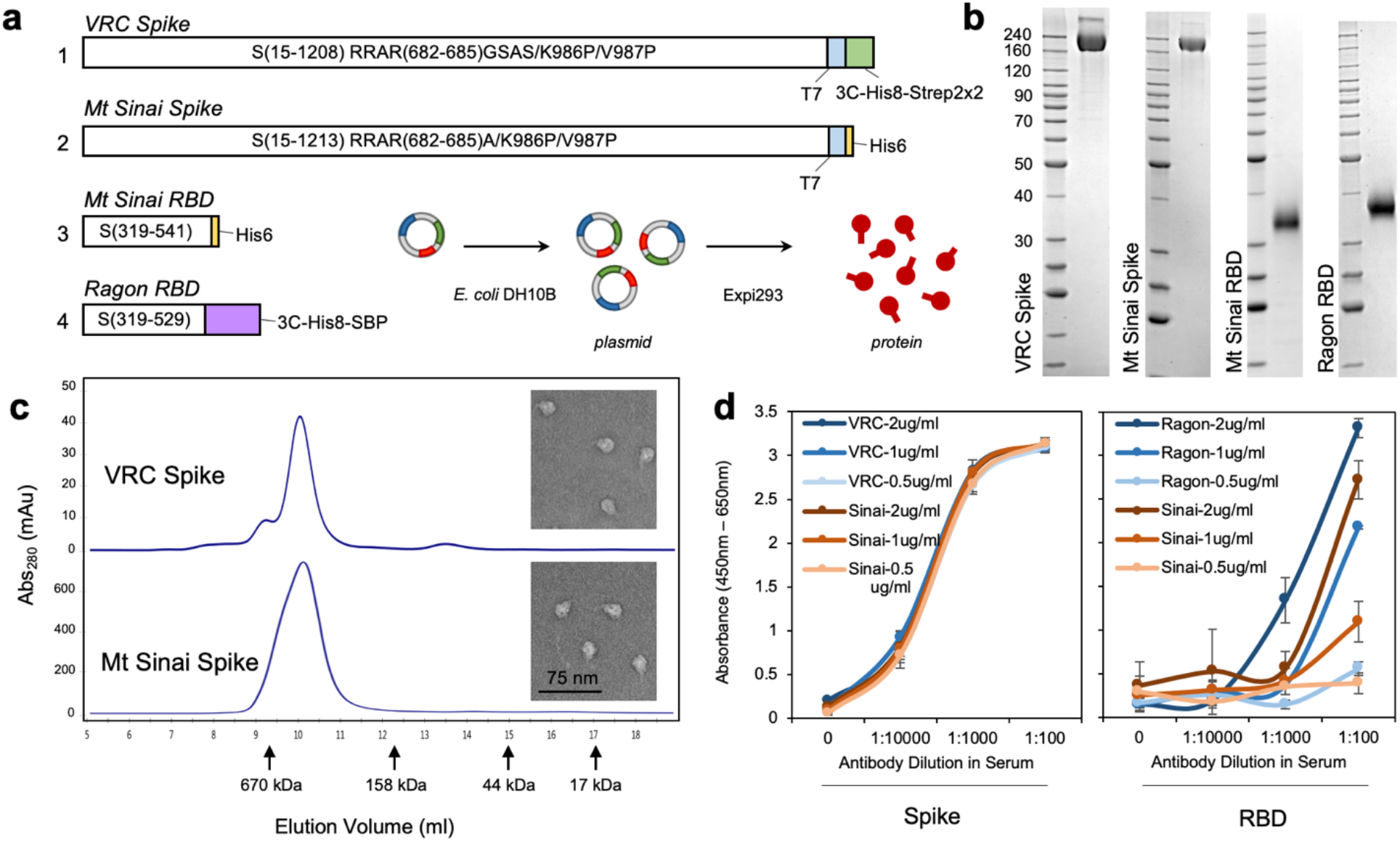
Production and sensitivity of two full spike ectodomain (Spike) and two receptor binding domain (RBD) constructs used as antigens in ELISA. **(a)** Schematic of the spike and RBD constructs used to generate recombinant proteins. Abbreviations are 3C, rhinovirus 3C protease cleavage site; Strep2x2, dual Strep2 epitope tag; T7, bacteriophage T7 fibritin trimerization domain; SBP, streptavidin-binding peptide. **(b)** SDS-PAGE Coomassie Blue staining of purified (1) VRC spike, (2) Mt Sinai spike, (3) Mt Sinai RBD, and (4) Ragon RBD proteins. **(c)** Analytical size exclusion chromatography of purified VRC and Mt Sinai spike proteins. Peak elution volumes of sizing standards are noted (670 kDa – thyroglobulin, 158 kDa – gamma-globulin, 44 kDa - ovalbumin, 17 kDa – myoglobin). Inset: transmission electron microscopy of VRC and Mt Sinai spike trimers. **(d)** Left: Full spike ectodomain at 3 different concentrations of protein coating density for both VRC (NIAID Vaccine Research Center, blue) and Sinai (Mount Sinai, orange) constructs. Right: RBD constructs at 3 different concentrations of protein coating for both Ragon (Ragon Institute, blue) and Sinai (Mount Sinai, orange) constructs. Anti-spike or anti-RBD monoclonal recombinant antibody spiked into negative serum at 1:100, 1:1000, and 1: 10000 dilution. Data are means ± SD. *n* = 4.

### Protein expression

Manufacturer’s protocols were followed for the transfection and culturing of Expi293 cells (Thermo Fisher Scientific, Waltham MA). After 96 hours (spike proteins) or 72 hours (RBD proteins) of expression, culture supernatants (8 liters for spike proteins and 4 liters for RBD proteins) were clarified by centrifugation (4000 x g, 20 minutes, 4°C) followed by filtration (Catalog# 12993, Pall Corporation, Port Washington, NY). Clarified supernatants were concentrated and buffer exchanged with the appropriate buffer by tangential flow filtration (TFF). Specifically, a MasterFlex peristaltic pump (Vernon Hills, IL) fed the clarified supernatant to either a 30 kDa MWCO cassette (Catalog# CDUF002LT, MilliporeSigma, Burlington, MA) for spike protein or 10 kDa MWCO cassette (Catalog# SK1P003W4, MilliporeSigma, Burlington, MA) for RBD. The clarified supernatant was concentrated to 10% of the initial volume and then buffer exchanged with 5 volumes of 1x PBS, pH 7.4 (Buffer A). Once the material was concentrated and buffer exchanged, the TFF cassette was rinsed with the appropriate buffer to collect any protein remaining in the cassette.

### Protein purification

All chromatography was conducted at room temperature (~22°C) using NGC medium-pressure chromatography systems from BioRad Laboratories Inc. (Hercules, CA). All spike proteins were purified similarly using immobilized metal affinity chromatography (IMAC) followed by desalting into final buffer. Specifically, TFF-treated culture supernatant was adjusted to 25 mM imidazole and applied to a 5 ml Ni Sepharose High Performance nickel-charged column (GE Healthcare, Chicago, IL), previously equilibrated in Buffer A amended to 25 mM imidazole. The flow rate for all steps of the IMAC was 5 ml/min. After the load, the column was washed in Buffer A + 25 mM imidazole for three column volumes (CVs) and the protein eluted from the column by first reversing the orientation of the column, then by applying increasing concentrations of imidazole (beginning with 75 mM and increasing each step by 50 mM ending with 475 mM) in Buffer A. Elution fractions were analyzed by SDS-PAGE and Coomassie-staining and appropriate fractions were pooled. The sample was desalted into the final buffer of 1x PBS, pH 7.4, (10X PBS 70011069 Thermo Fisher Scientific, Waltham, MA) using a HiPrep 53 ml 26/10 desalting column (GE Healthcare, Chicago, IL) with 14 ml injections at 9 ml/min for all steps. The final protein sample was created by combining the bulk elutions from multiple runs of the desalting column. The protein concentration was determined by measuring the A280 using a Nanodrop One spectrophotometer (Thermo Fisher Scientific, MA, USA). Final protein was dispensed as 0.5 ml and 0.05 ml aliquots, snap frozen in liquid nitrogen, and stored at −80°C. One ml of the final protein was thawed and analyzed by analytical size-exclusion chromatography with a 10/300 Superdex200 analytical column (GE Healthcare, Chicago, IL), using a flow rate of 0.5 ml/min, to confirm the trimeric nature of the protein. Transmission electron microscopy of the purified VRC and Mt. Sinai spike proteins was also carried out by dilution of the samples to 0.02 mg/mL in 20 mM Tris-HCl, pH 8.0, 200 mM NaCl followed by loading onto glow-discharged carbon film grids. Grids were washed twice in buffer, stained with 0.75% w/v uranyl formate (pH 4.5) four times, and stain was then wicked off the grid and grids were dried under an incandescent lamp. Stained grids were imaged on a Hitachi 7650 electron microscope at 40,000x magnification.

Mt. Sinai RBD and Ragon RBD proteins were purified similarly using IMAC followed by size exclusion chromatography (SEC) into final buffer using NGC medium-pressure chromatography systems from BioRad Laboratories Inc. (Hercules, CA). Specifically, TFF-treated culture supernatant was adjusted to 25 mM imidazole and applied to a 10 ml Ni Sepharose High Performance nickel-charged column (GE Healthcare, Chicago, IL), previously equilibrated in 1x PBS, pH 7.4, amended to 25 mM imidazole. The flow rate for all steps of the IMAC was 5 ml/min. The column was washed in 1x PBS, pH 7.4, 25 mM imidazole for four CVs. Proteins were eluted from the column with a gradient of 1x PBS, pH 7.4, from 25 mM to 500 mM imidazole over 20 CVs. Elution fractions were analyzed by SDS-PAGE and Coomassie-staining and appropriate fractions were pooled. The sample (~40-50 ml) was concentrated to ~5 ml using 10 kDa MWCO Amicon Ultra centrifugation filter units (MilliporeSigma, Burlington, MA), and the sample applied to a 16/60 Superdex75 SEC column (GE Healthcare, Chicago, IL) equilibrated in final buffer of 1x PBS, pH 7.4. The column was developed at 1 ml/min and one ml fractions were collected. Fractions were analyzed by SDS-PAGE and Coomassie-staining and appropriate fractions were pooled and filtered with a 0.22 μM syringe filter (low protein binding). The protein concentration was determined by measuring A_280_ using a Nanodrop One spectrophotometer (Thermo Fisher Scientific, MA, USA). Final protein was dispensed as 0.25 ml and 0.05 ml aliquots, snap frozen in liquid nitrogen, and stored at −80°C.

### Enzyme-linked Immunosorbent Assay

Serum samples collected via venipuncture in serum separator tubes were processed and stored in 200ul aliquots at −80°C before analysis. Whole blood samples were loaded onto 20 μl Neoteryx Mitra Microsampling device tips, dried completely, and transferred into 500 μl Eppendorf tubes that were stored dry at −80°C until elution. Prior to analysis via ELISA, one microsampler tip (20 μl) was added to 400 μl of 1XPBS (Gibco) + 1.0% BSA (Sigma) + 0.5% Tween20 (Sigma) in a 1 ml deep-well 96 well plate (ThermoFisher). The plate was then covered with an adhesive foil seal and incubated overnight at 4°C on a shaker at 300 rpm. The resulting eluate was used immediately for ELISA or stored at −80°C until use.

One hundred (100) microliters of spike (1 μg/ml) or RBD (2 μg/ml) antigen was added into 96-well Nunc MaxiSorp ELISA plates (ThermoFisher) and incubated for 16 hours at 4°C. Wells were washed with 300 ul of 1xPBS + 0.05% Tween20 (PBS-T) three times and blocked with 200 μL PBS-T + 5.0% Nonfat Dry Milk (blocking buffer) for 2 hours at room temperature. The plate was then washed three times with 300 μl PBST. One hundred microliters of each sample were added in technical duplicate (serum diluted 1:400 in blocking buffer, microsampler eluate diluted 1:10 in 1xPBS + 5.0% Non Fat Dry Milk) to the plate. Controls on each plate included blank/secondary only control, known nasal swab positive serum control, archival serum negative control, and recombinant antibody positive controls (NIAID VRC). Samples were incubated at room temperature for 1 hour, and plates were washed three times with 300 μl PBST. Secondary antibody (HRP conjugated: Goat anti-Human IgG (H+L) Cross-Adsorbed Secondary Antibody, Goat anti-Human IgM Cross-Adsorbed Secondary Antibody, Goat anti-Human IgA Cross-Adsorbed Secondary Antibody; ThermoFisher) was diluted at 1:4000 in blocking buffer and 100 μl of each antibody was then added to each well and incubated for 1 hour. Plates were washed three times with PBST, then incubated with 1-Step™ Ultra TMB-ELISA Substrate Solution (ThermoFisher) for 10 minutes prior to stopping the reaction with 1 N sulfuric acid Stop Solution (ThermoFisher). Plates were then read for absorbance at 450 and 650 nm (PHERAstar FSX plate reader) within 30 minutes of stopping the reaction. Absorbance (OD) is calculated as the absorbance at 450 nm minus the absorbance at 650 nm to remove background prior to statistical analysis.

### Defining Thresholds for Positivity

To evaluate the specificity of ELISA assays and establish thresholds for positivity, 100 serum samples collected from well-characterized healthy volunteers in NIH study NCT01386424 prior to 2019 were obtained as negative controls for SARS-CoV-2 to define the threshold for seropostivity and evaluate specificity. A total of 14 convalescent blood samples from known SARS-CoV-2 nasal swab PCR+ donors were obtained to evaluate sensitivity. A test sample set for use to further assess the overall serologic assays testing protocol was also obtained. This set of deidentified blood samples (no donor data associated with sample) was acquired from a blood drive in very high-risk Jewish communities in New York and New Jersey, of which 22 were from PCR+ diagnosed individuals, while 46 were from individuals with a high chance of exposure as well as reported symptoms, while 6 donors displayed no symptoms but had a high chance of exposure. These samples were tested for the presence of SARS-CoV-2 antibody and were also run against a panel of spike proteins from four other beta-coronaviruses; MERS, SARS1, OC43 and HKU1 to establish seroprevalence estimates for these pre-pandemic coronaviruses in a current population.

### Statistical Analysis and Determination of Sensitivity and Specificity

To determine confidence intervals for specificity and sensitivity, we used exact binomial methods, which to give accurate coverage require an independent data set to determine the threshold. Because we used the same data set to determine the threshold and evaluate sensitivity and specificity, we only considered a restricted class of thresholds, determined by adding an integer values of standard deviation to the mean for the negative controls. This approximates accurate confidence intervals. Using this procedure and the OD responses from testing the 100 negative control samples on both the SARS-CoV-2 spike and RBD assays, thresholds for each of the 4 primary types of antibody OD responses measured (spike IgG, spike IgM, RBD IgG or RBD IgM) were used to give a tentative threshold. Further negative control samples evaluated in the future will be collected and evaluated so that the lower 95% confidence limit is greater than 99%.

To evaluate how well our final testing protocol would work in practice, simulation studies were performed to determine how the number of samples used to estimate sensitivity and specificity affect the confidence interval of the prevalence in a serosurvey. Several scenarios were simulated to evaluate the performance of the estimation methods with simple weighting and different sample sizes used to estimate the sensitivity and specificity. In the simulations, data were generated from a survey size of 10,000 under overall true prevalence rates of 0.1%, 1%, and 10%. Scenarios with two different sets of observed weights were evaluated, first with samples drawn from two populations with the true proportion of people in each population being 0.35 and 0.65, but were drawn equally from each population. The weights for each person in the smaller and larger populations were approximately 0.7 and 1.3 respectively. In the population with the smaller size the true prevalence rate was inflated above the overall rate, while the true prevalence in the second population was below the overall rate. A second set of more extreme weights was examined. In this simulation the proportion in each population were 5% and 95% which corresponded to weights of 0.1 and 1.9 respectively.

We performed simulations with true specificity values of 99% and 100%, using sample sizes of 100, 300 and 1000 to estimate the specificity. A true sensitivity of 90% was used in these simulations with a sample size of 100 used for the estimate. Another set of simulations was performed with true sensitivity values of 90% and 95% and sample sizes of 100, 200, and 300 used in the estimate with true specificity set to 99% with sample size of 1000 used in the estimate. We performed 1000 simulations for each scenario and calculated 95% confidence intervals around the adjusted prevalence estimate for each replication of the simulation.

## RESULTS

### Optimization of protein production and purification

The CoV-2 spike protein receptor-binding domains (RBD) were produced by secretion from Expi293 cells at high yield (>15 mg/l) after 72 hours of expression. Little improvement in yield was seen with extended expression times. Although the Mt. Sinai and Ragon constructs differ significantly in their C-terminal tags and use different secretion signal peptides (native spike for Mt. Sinai, tissue plasminogen activator (TPA) for Ragon), the behavior of these proteins during purification was highly similar. SARS-CoV-2 spike proteins, in contrast, were poorly secreted with yields of no greater than 2 mg/l in our hands, similar to reports in the literature^8,9^. Improved production was seen at 96 hours versus 72 hours of expression; however, viabilities of cultures were very low after 96 hours, suggesting toxicity perhaps due to failures in the secretory pathway or activation of the unfolded protein response. Curiously, expression of other coronavirus spike proteins (SARS1, MERS, OC43, HKU1) consistently produced higher yields (5-11 mg/l) compared with CoV-2 for reasons that are still unclear. Purification of spike proteins was further complicated by significant protein loss during size exclusion chromatography. Ultimately, we chose to eliminate the SEC step and instead modified the IMAC purification conditions to produce higher purity material, followed by desalting to final buffer. This eliminated the protein loss during SEC and produced proteins that were similar in purity to those seen with the original protocol. **Figure 2b** shows the final purified proteins generated using the modified protocols. To verify that spike proteins formed the expected trimeric structures, analytical size exclusion chromatography was performed (**Figure 2c**) and VRC and Mt. Sinai spike proteins showed a clear elution peak corresponding to the size of a trimer, with no apparent monomeric peaks. In addition, transmission electron microscopy images (**Figure 2c, inset**) confirmed the presence of particles similar in size and appearance to previously characterized CoV-2 trimers^9^. Taken together, these data suggest that the highly pure spike proteins are properly folded into their expected trimeric structures. Very little difference was observed in the purity or yield of the VRC and Mt. Sinai spike proteins, which is not surprising given the similarity in their protein sequences.

### Technical performance of multiple antigen constructs

In order to determine the optimal assays for use in our serology protocol for SARS-CoV-2, we compared multiple antigens currently in use. These constructs included the spike ectodomain constructs from the NIAID/VRC and from Florian Krammer’s laboratory at Mount Sinai, as well as one RBD construct from the Ragon Institute of MGH, MIT and Harvard, and one from the Krammer Lab of Mount Sinai^8,9^ (**Figure 2d**). Both spike constructs behaved similarly and showed no difference in ELISA signal across a range of recombinant antibody dilutions in serum and blood that was loaded onto microsamplers. They also displayed similar sensitivity at multiple coating densities from 0.5 – 2 μg/ml. The two RBD constructs were similarly compared and displayed significantly different sensitivity at a range of recombinant antibody concentrations and coating densities. The RBD construct from the Ragon Institute displayed stronger signal with a recombinant RBD antibody when compared to the Krammer Lab construct. Therefore, based on these data we chose to use the VRC spike construct and Ragon institute RBD for our serological protocol by testing samples for the presence of antibody against both constructs.

### Sensitivity and Specificity of Selected ELISA Assays and Final Protocol

To evaluate specificity of the assays, 100 negative controls (serum collected ‘pre-2019’ and therefore prior to the emergence of SARS-CoV-2, **Figure 3**) were assessed. At a 1:100 dilution of serum in the SARS-CoV-2 spike ELISA, there was an unacceptable amount of background signal from the negative controls (**Supplementary Figure 1**). At a 1:400 dilution, no loss of signal was observed from the positive controls and background was significantly diminished when assessing the negatives. At a threshold of 2 standard deviations above the mean of the negative controls, estimated 98% specificity was noted on the spike assay, while at a threshold of 3 standard deviations above the mean (OD = 1.078), estimated 99% specificity was observed (**Figure 3**). We repeated this assay for both IgM (OD = 0.405) and IgA (OD = 0.308), which resulted in estimated 100% and 99% specificity at 3 standard deviations above the mean. This was repeated with the RBD construct, where we observed 100% specificity for IgG (0.264), IgM (OD = 0.213), and IgA (OD = 0.723) at 3 standard deviations above the mean of the negative controls. Based on these data we propose using a threshold of 3 standard deviations above the mean of the negative controls for both the spike and RBD manual ELISA assays. Therefore the definition of seropositivity in our protocol requires positivity (above the threshold) of both spike IgG and RBD IgG or both spike IgM and RBD IgM. This definition provides a specificity of 100% (95% CI 96.4%, 100%) from our negative controls. With the accrual of further negative controls this can be fine tuned to thresholds to ensure specificity with a 95% lower confidence limit of at greater 99%. We are utilizing IgM and IgG to threshold seropositivity as early infection could be signaled with IgM+ but IgG-donors; however, we expect the majority of seropositive individuals to be IgG+.

**Figure 3:**
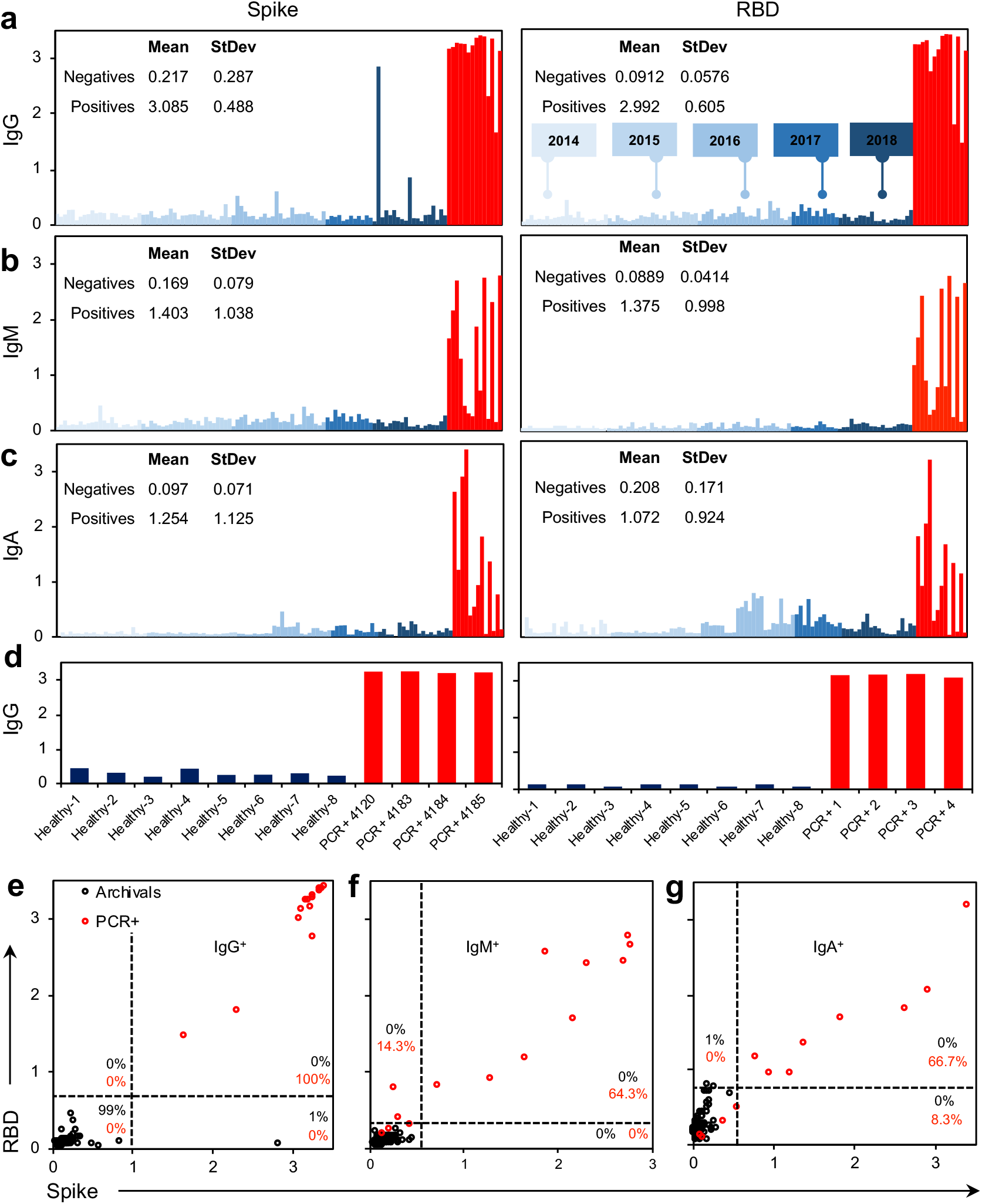
Specificity of antigens against a preliminary panel of pre-2019 archival sample controls. **(a-c)** Spike (VRC) and RBD (Ragon) antigens tested against 100 archival controls collected from 2014 - 2018 (blue) compared with 14 SARS-CoV-2 PCR+ controls (red) for **(a)** IgG, **(b)** IgM, and **(c)** IgA. **(d)** Microsampler controls of PCR+ samples against healthy controls, **(e-**f) Seropositivity thresholding for (e) IgG, (f) IgM) and (g) IgA calculated from average signal intensity of archival controls, *n =* 100 archival controls, *n =* 14 nasal swab SARS-CoV-2 PCR+ patients. Manual ELISA, titer = 1:400 (serum) or 1:10 (microsampler eluate).

Sensitivity was evaluated by testing the 14 positive control samples (i.e. serum collected from convalescent patients previously confirmed by PCR diagnostic testing with SARS-CoV-2). Samples were tested at the optimal 1:400 dilution observed for manual ELISA. Using our previously defined thresholds of 3 standard deviations above the mean of the negative controls for both spike and RBD, all positive control samples met the criteria for positivity. Therefore, the preliminary estimate of sensitivity is 100% (95% CI 76.8%, 100%) based on a small sample size. An increased sample size of positive controls added in the future will allow for further tightening of this confidence interval and a more accurate measure of sensitivity.

Additionally, blood loaded onto the microsamplers and dried and stored prior to elution was compared to standard frozen serum. Antibodies reliably eluted from the microsamplers and were detected. Subsequently antibody titers were performed and reliable titration of both IgG and IgM antibodies from positive control blood was achieved. Furthermore, when comparing 68 serum and microsamplers from the same donors, estimating a 50% serum volume in blood, there is a strong correlation between microsampler OD and serum OD (R^2^ = 0.983 for IgG, R^2^ = 0.923 for IgM) at a 1:1 ratio (slope = 1.0015 for IgG and 0.9569 for IgM). This suggests that not only could detection of antibodies be performed reliably, but semi-quantifying antibodies from these mail-in sampling devices was possible (**Supplementary Figure 2**).

### Results from a Small-Scale Test Sample Set

In order to further evaluate this serology testing protocol, we analyzed a set of samples from a high-exposure community. The PCR status and symptom status of the individuals were not linked to each sample, but all 68 donors were known to have exposure to COVID19, 22 of whom had also tested positive in the recent past for SARS-CoV-2 infection by PCR. Using our protocol of testing we identified 59 individuals (from a total of 68) who met our criteria for positivity with both spike and RBD above the threshold for IgG and IgM. Subdividing the results based on the individual assays themselves, 59 tested positive for spike IgG, 31 tested positive for spike IgM, 62 tested positive for RBD IgG, and 51 tested positive for RBD IgM (**Figure 4a-d**). When directly comparing the OD of spike versus RBD, we saw a correlation for IgG (R^2^ = 0.856) and IgM (R^2^ = 0.961) (**Figure 4b,d, Supplementary Figure 3**). The correlation of IgG was less than for IgM, and several points fell below the trendline when plotting spike as a function of RBD, suggesting that using spike alone risks overestimating overall intensity of SARS-CoV-2 antibody signal, which is predictable given the increased availability of antigenic sites on the full spike ectodomain and possible cross reactivity (**Supplementary Figure 3b**). Including RBD in the protocol offers increased specificity reducing the risk of false positives, while including spike offers increased sensitivity able to capture a polyclonal response. Therefore, using these antigens individually risks false results, while together they provide the desired sensitivity and high specificity. Overall, we identified 37 undiagnosed COVID19 cases in this sample dataset (spike IgG+, RBD IgG+). Furthermore, as asymptomatic transmission has been reported^10^, we tested six additional samples that were sourced from donors from this high-exposure community that displayed no symptoms, and were able to detect antibody in four of the six samples tested (**Figure 4f**).

**Figure 4:**
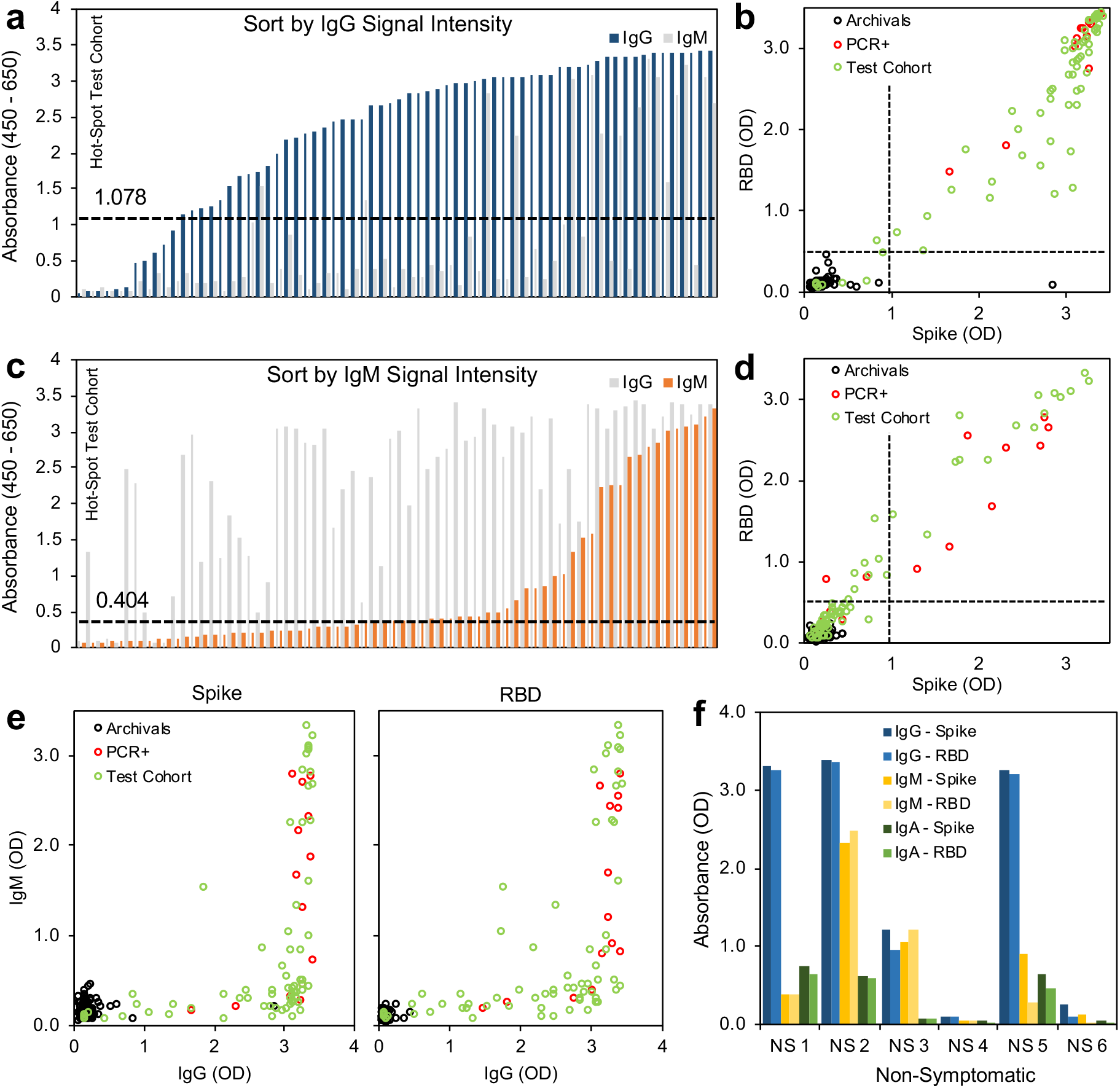
Small-scale testing of SARS-CoV-2 seropositivity from a hard-hit community. **(a)** Signal intensity (absorbance) sorted by IgG **(b)** Thresholding of seropositivity in small-scale test cohort for SARS-CoV-2 IgG, black = archival negative controls, red = known PCR-diagnosed positive controls, green = test cohort, **(c)** Signal intensity (absorbance) sorted by IgM. **(d)** Thresholding of seropositivity in small-scale test cohort for SARS-CoV-2 IgM **(e)** Relation between IgM expression and IgG expression of spike and RBD antigens, **(f)** Seropositivity in non-symptomatic individuals that have not tested (PCR) positive for SARS-CoV-2 infection show robust IgG expression in absence of symptoms, *n =* 68 symptomatic donors, *n =* 6 non-symptomatic donors.

### Cross-reactivity of SARS-CoV-2 antibodies with other beta-coronaviruses and respiratory viruses

To evaluate the potential for cross-reactivity altering any ELISA results^11,12^, all control samples were tested for the presence of antibody against the spike antigens of OC43, HKU1, MERS, and SARS1. We also tested all of the samples from our test sample set for these antibodies as well. When evaluating archival negative controls, we had previously shown very low signal for SARS-CoV-2 spike ELISA (negative, **Figure 3a**). When compared with spike proteins from other coronaviruses, we detected very high OC43 and HKU 1 antibodies across the pre-2019 sample set, concluding that there is minimal cross-reactivity between OC43 and HKU1 antibodies and the SARS-CoV-2 spike protein. Both OC43 and HKU1 spike antigens resulted in high absorbance readings in the test samples set (3.07 ± 0.22 and 2.49 ± 0.62), SARS-CoV-2 positive controls (3.25 ± 0.16 and 2.92 ± 0.49), and SARS-CoV-2 negative controls (2.93 ± 0.47 and 2.30 ± 0.65), suggesting a strong prevalence of seasonal coronavirus infection that limits ability to assess cross-reactivity due to high frequency of standard seasonal coronavirus infection in the general population (**Figure 5a, Supplementary Figure 4**). SARS1 and MERS spike antigen ELISAs resulted in significantly lower signal intensity when compared to SARS-CoV-2 (SARS-CoV-2 = 2.58 ± 1.00, SARS1 = 0.93 ± 0.86, MERS = 1.09 ± 0.89). However, as both proteins did induce higher than background levels of absorbance, we evaluated their expression on the negative controls. Overall no correlative effect (R^2^ = 0.3577, 0.2606, 0.2747, and 0.4174, respectively) was found between SARS-CoV-2 signal intensity and the signal intensity of the four other coronaviruses (OC43, HKU1, MERS, SARS1). We also found no correlation between the hemagglutinin inhibition titer to H1N1 influenza A and the SARS-CoV-2 signal intensity, suggesting negligible cross-reactivity between antibodies against other common respiratory viruses and the antigens used in our assays (**Supplementary Figure 5**),

**Figure 5:**
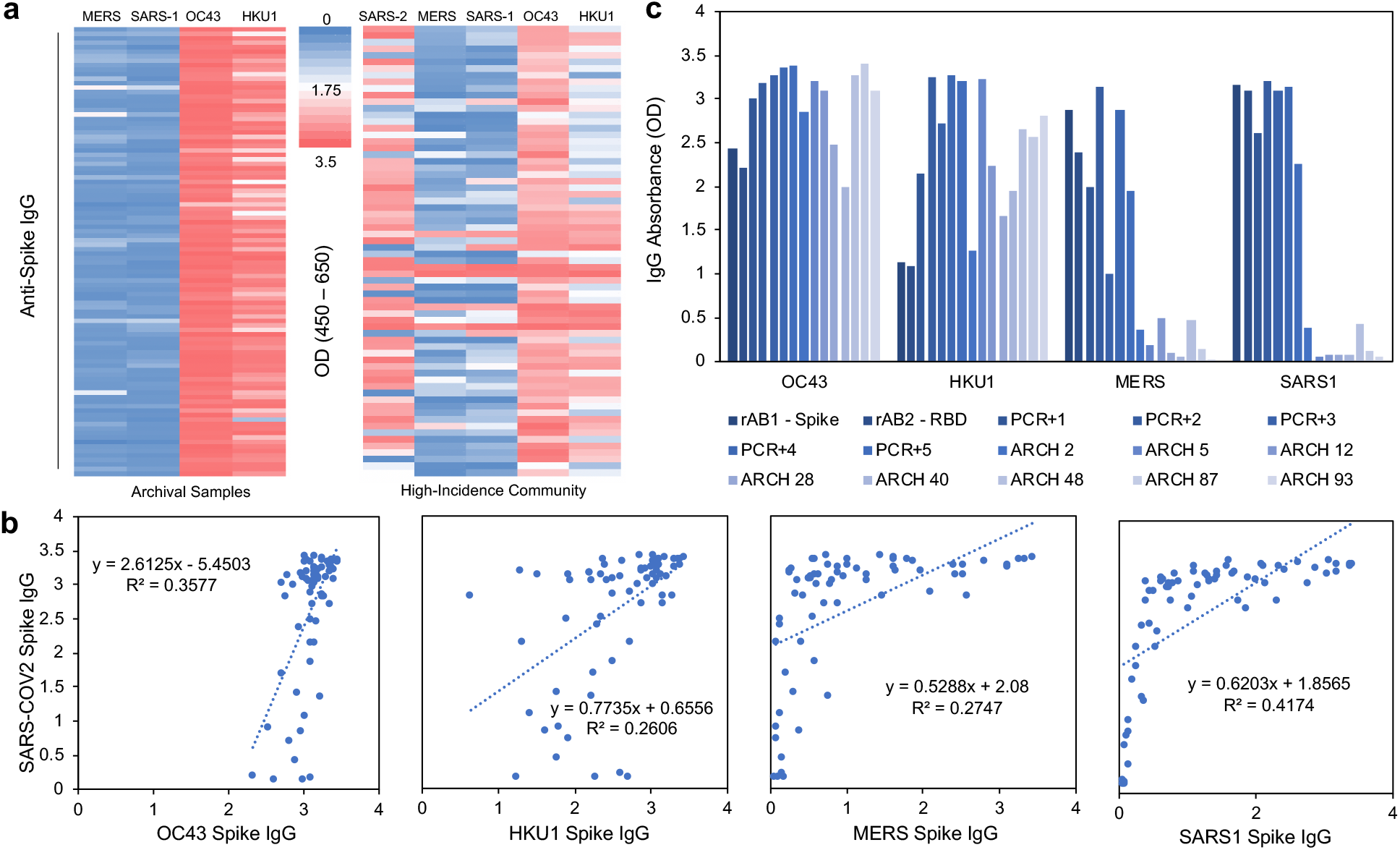
Cross-evaluation of high seroprevalence community samples with spike antigens from other coronaviruses. **(a)** IgG absorbance of spike antigens from SARS-CoV-2, previous epidemic (MERS and SARS1), and seasonal (OC43 and HKU1) coronaviruses in archival pre-2019 samples (left) and high-incidence community (right), **(b)** Scatter plots (with linear models and R-squared values) between IgG signal from SARS-CoV-2 spike and other coronavirus spike proteins, **(c)** Signal intensity for recombinant antibody control (rABl = spike monoclonal; rAB2 = RBD monoclonal), known SARS-CoV-2 nasal swab positive patient control (PCR+), and pre-2019 archival control (ARCH) samples for OC43, HKU1, MERS and SARS1.

### High-Throughput Automation and Reproducibility of Results

To increase our throughput and scale our studies, we moved toward a robotic high-throughput semiautomated platform. In addition to increasing the speed of sample processing, automation also increases standardization, minimizes technical variability, and keeps protocols consistent over long periods of time and multiple operators. As such, we implemented a parallel plate washer/dispenser in line with automated plate stackers and plate reader (**Supplementary Figure 6**).

Utilization of this platform minimized well-to-well and plate-to-plate technical variability (**Supplementary Figure 7**). As automated systems alter the conditions and timing and do not precisely replicate manual pipetting, we re-titered known PCR+ patient positive controls (*n* = 9) and archival negative controls (*n* = 8) representing the range of signal intensity (min to max) observed with both positives and negatives detected by the manual ELISA (**Supplementary Figure 8**). Given the biologic difference in the concentrations of IgG, IgM and IgA, and based on the balance between background signal and positive signal, we have found that a 1:1000 to 1:400 dilution for IgG results in clear signal with high positives (higher concentration in serum) and a 1:400 dilution for IgM and IgA analyses is necessary to capture all positives (lower concentration in serum). We re-validated these titers on sero-positive and sero-negative identified donors from our small-scale test-cohort using microsamplers loaded with donor blood then eluted as per standard protocol (**Supplementary Figure 9**). To ensure equal endpoint serum dilution, we use an equivalent dilution of microsampler eluate for IgG analyses (1:25 to 1:10, corresponding to 1:1000 to 1:400 serum dilution), and 1:10 for IgM and IgA analyses, equal to a 1:1000 and 1:400 serum dilution, respectively. To re-establish thresholds and sensitivity on our automated setup that will be used in future serosurvey work, we evaluated the robotic setup against 246 archival pre-2019 negative controls (**Supplementary Figure 10, 11**). At a threshold of the mean of the archival controls plus three standard deviations we detect 0 false positives for IgG (thresholds: spike = 0.669, RBD = 0.432) and 1 false positive for IgM (thresholds: spike = 0.271, RBD = 0.277). Therefore the preliminary specificty for IgG is 100% (95% CI 98.5, 100.0) and the for IgM preliminary specificity of 99.59% (95% CI 97.8, 100.0). If we increase the threshold for IgM to 4 standard deviations above the mean, we detect 0 false positives giving us a 100% preliminary specificity of 100% (95% CI 98.5-100). Further positive and negative controls will be assessed to confirm titers, and further define the appropriate thresholds and final sensitivity and specificity confideince intervals when using the automated system.

### Statistical Simulation of Confidence Intervals in Relation to Validation of Assays

To evaluate our assay in the context of future serosurvey efforts, we modeled the statistical confidence over a range of disease prevalence and assay specificity. We simulated a study with 10,000 subjects and two strata with different prevalence under different scenarios changing the stratum-specific prevalences, the true sensitivity, true specificity, negative control and positive control sample sizes. In all simulations the confidence intervals included the true value over 95% of the replications. **Figure 6** shows the plot of the lower and upper 95% confidence interval for the prevalence for each of the 1000 replications for each scenario. The supplemental materials provide the equations used to estimate the prevalence and 95% confidence intervals adjusted for weighting and estimated sensitivity and specificity. The confidence intervals are sorted by the lower confidence interval. The figure shows that true specificity of 100% dramatically improves the width of the confidence intervals for all true prevalence rates. The figures also show an improvement as the negative control sample size increases from 100 to 300 and then 1000. This improvement is especially important in a low true prevalence scenario. The simulations with prevalence .001 (to simulate a true prevalence of near 0) shows that the lower bound will be 0 with a sample size of at least 300 negative controls to estimate specificity, and most upper bounds will be less than 1%. The simulations with prevalence of 1% again show the importance of a large sample size to estimate specificity. For negative control sample sizes of 1000 and a specificity of 1, the lower estimate is above 0 and the upper is almost always less than 1.5%. This will provide a very tight confidence interval in estimating the prevalence. These simulations show the importance of having high specificity, and if the expected prevalence rates are low, the importance of basing specificity estimates on large negative control sample sizes. When simulating less extreme weighting situation, the simulation with the more extreme weights generates very similar results (results not shown).

**Figure 6:**
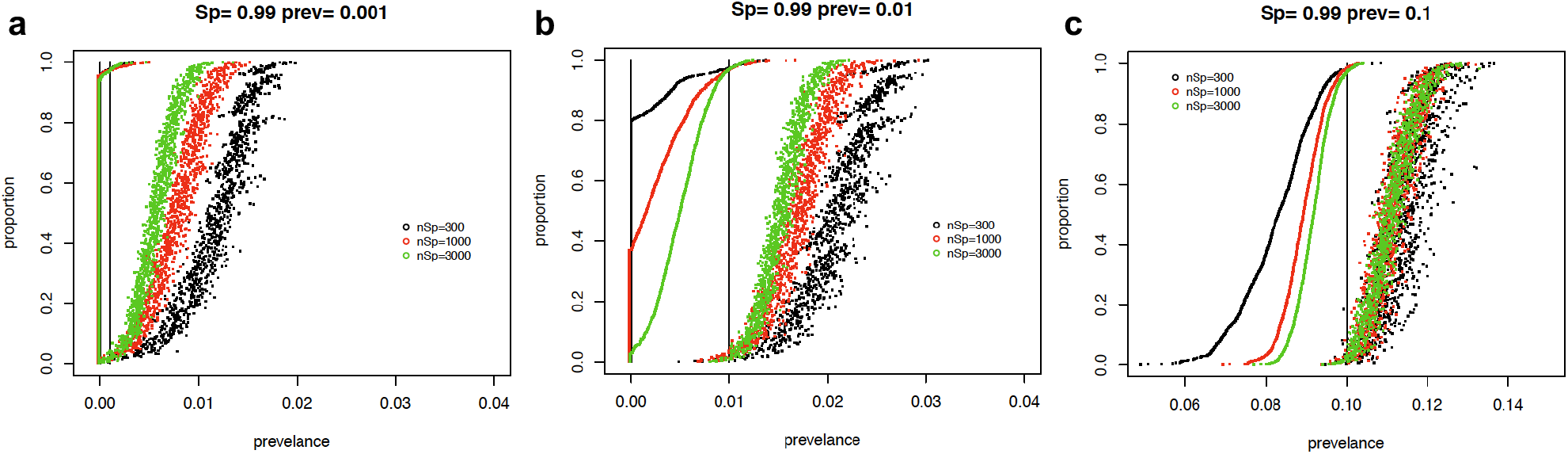
Simulation results showing confidence intervals for serosurvey prevalence calculations. Each graph displays 95% confidence intervals (CI) from 1000 replications of each condition. The CIs are sorted by the lower bound. For graphs a-c the true sensitivity is 0.90 and it is estimated with 100 samples. The true specificity is 0.99 and it is estimated with sample sizes of 100 (black), 300 (red), 1000 (green). Each graph shows the different underlying true prevalence values of either 0.001, 0.01, or 0.1. The true prevalence for each simulation is given by a vertical line.

We further analyzed the impact of different sensitivity values and different sample sizes to estimate the sensitivity on the width of the 95% confidence intervals for the estimate of prevalence (**Supplementary Figure 12**). This figure shows that there is minimal effect on the intervals for increasing positive control sample size above 100 and increasing sensitivity above 90%.

## DISCUSSION

Understanding the seroprevalence of SARS-CoV-2 antibodies in the general population is critical to understanding the extent of the spread of SARS-CoV-2 infection. However, design of assays and evaluation of seroprevalence data is not trivial. Due to the variable nature of the human species, the technical intricacies of assay development, the potential impact of seroprevalence measurements on policy adjustment, and the current uncertainty of the correlation between antibody presence and immunity, these assays and trials must be approached with caution and rigor^13^.

In this study, we compared the activity of four different constructs used in ELISA for serology of SARS-CoV-2 and established the ELISA assay being utilized in our seroprevalence study. First, we developed optimized protein expression and purification methods that could reproducibly generate spike and RBD proteins of consistent yield and quality. While RBD production is fairly straightforward, production of properly folded trimeric spike proteins is considerably more challenging. Spike proteins, likely due to their heavy glycosylation, have a tendency to adsorb to membranes as well as purification media, requiring more complex methods to ensure consistent yield and quality. After experiencing significant protein loss during size exclusion chromatography using multiple purification resins, we decided to forgo this step and alter the preceding IMAC chromatography with a more complex elution scheme that minimized levels of contaminants and retained protein purity. This modified procedure has proved more consistent and allowed us to generate single large batches (>15 mg) of spike protein from 8 liters of cell culture media permitting thousands of assays to be run on a single qualified batch of protein. Attention to quality of protein production and purification is critical for running a validated and specific ELISA, as contaminants in impure protein preparations or unstable proteins can yield decreased specificity and sensitivity in the assay.

Through careful evaluation of various ELISA assays and statistical determination of optimal threshold cutoffs for specificity, we determined that a combination approach using two ELISA assays, one employing the VRC spike construct and the other employing the Ragon RBD, provided optimal results (Figure 6). To be scored as ‘positive’, both the spike IgG and RBD IgG OD levels or both the spike IgM and RBD IgM levels must be above their respective thresholds. Based on the data presented here, this preliminary method provides an estimated sensitivity of 100% (95% CI: 76.8%, 100%) and specificity of 100% (95% CI: 96.4%, 100%) when using both for IgG and IgM. As we deploy this method for a large-scale NIH serosurvey study, we will continue to test control cases to achieve a minimum of 300 and up to 1000 negative controls, and a minimum of 100 or more positive controls. As this is completed the thresholds can be further updated to ensure that the 95% lower confidence limit on specificity is greater than 98%. Final thresholds can then be determined and applied in future studies. In addition to the adjustment of these thresholds, estimates of prevalence from serosurveys using this method may use weighting methods (e.g., propensity weighting) and should adjust for the specificity and sensitivity estimates and variability about these estimates. The supplemental materials provide the equations that can be used to adjust the observed prevalence estimates based on the number of control samples tested.

The adaptation of this protocol to analyze both IgM and IgA allows characterization of the stage of infection and increases the sensitivity to identify relatively early-stage infections that have yet to mount a strong IgG response. This will allow for additional research to be performed during serosurveys, prospective, or larger-scale testing to better understand the development of immunity and the timing of various antibody responses.

The analysis of data from the small sample set collected from communities with high transmission rates in New York City and New Jersey demonstrated how people in various stages of their antibody responses may appear in our assays. There were 68 donors known to have exposure to and reported symptoms of COVID19, 22 of whom had also tested positive in the recent past for SARS-CoV-2 infection by PCR. Using our protocol, we identified 86% of these symptomatic and highly exposed individuals which were seropositive for IgG. We also identified 31 who were IgM positive while also being IgG positive (suggesting they were in an early stage of recovery from disease), while others were solely IgG positive with IgM below the positive thresholds for spike and RBD (suggesting a later stage of convalescence)^14^. Overall expression of RBD and spike correlated well for both IgG and IgM, but several mid-range donors displayed lower RBD absorbance levels (still within positive range) when compared to spike, suggesting these individuals may have a polyclonal antibody response that is not captured by the RBD antigen alone.

It is important to note that we developed this protocol with ease of adaptation by other labs in mind, and we focused on utilizing readily available reagents and instruments so that it could be applied easily in various resource settings. However, appropriate validation must be performed at each lab that adopts this protocol due to variances in equipment and reagents. These validations include determining the proper dilutions and building confidence intervals with positive and negative sample controls. This protocol, if validated and applied properly, can provide sensitive and highly specific data that are more reliable than those of binary threshold assays and can serve to develop a more complete understanding of humoral immunity. With further validation, this protocol will be implemented in our current NIH serosurvey and we believe that it could offer a consistent method for others performing similar studies or expanded clinical antibody testing in the future.

## Data Availability

The data contained in this manuscript will be available at request after formal publication of the manuscript in a peer-reviewed journal.

## ACKNOWLEDGEMENTS

The authors would like to acknowledge Kizzmekia Corbett and Barney Graham of the NIAID VRC for their generous donation of coronavirus spike plasmids and recombinant antibodies, and Dr. Aaron Schmidt, Jared Feldman, Blake M. Hauser and Timothy M. Caradonna for their donation of their RBD expression plasmid. We would like to thank Golan Ben-Oni, Rabbi Shua Brook, Dr. Adam Polinger, Dr. Avi Rosenberg, and the Jewish community of New York and New Jersey for their generous donation of blood samples used to validate and test this assay. This research was supported in part by the Intramural Research Program of the NIH, including the National Institute for Biomedical Imaging and Bioengineering, the National Institute of Allergy and Infectious Disease, and the National Center for Advancing Translational Sciences. This project has been funded in part with Federal funds from the National Cancer Institute, National Institutes of Health, under contract number HHSN261200800001E. <u>Disclaimer:</u> The NIH, its officers, and employees do no recommend or endorse any company, product, or service.

## AUTHOR PREPRINT DISCLAIMER

*This is a non-peer reviewed pre-print manuscript representing a work that has been released due to pertinence to the current SARS-CoV-2 public health crisis. Rapid release of information is necessary; however, it does not represent the full extent of final validation for the associated serosurvey*.

